# Validation of Visual and Auditory Digital Markers of Suicidality in Acutely Suicidal Psychiatric In-Patients

**DOI:** 10.1101/2020.10.21.20217091

**Authors:** Isaac R. Galatzer-Levy, Anzar Abbas, Anja Ries, Stephanie Homan, Vidya Koesmahargyo, Vijay Yadav, Michael Colla, Hanne Scheerer, Stefan Vetter, Erich Seifritz, Urte Scholz, Birgit Kleim

## Abstract

**Background:** Multiple symptoms of suicide risk are assessed based on visual and auditory information including flattened affect, reduced movement, and slowed speech. Objective quantification of such symptomatology from novel data sources can increase the sensitivity, scalability, and timeliness of suicide risk assessment.

**Methods:** In the current study we utilized video to quantify facial, vocal, and movement markers associated with mood, emotion, and motor functioning from a structured clinical conversation in 20 patients admitted to a psychiatric hospital following a suicide risk attempt. Measures were calculated using open source deep learning algorithms for processing facial expressivity, head movement, and vocal characteristics. Derived digital measures of flattened affect, reduced movement, and slowed speech were compared to suicide severity using the Beck Suicide Scale (BSS), controlling for age and gender using multiple linear regression.

**Results:** Suicide severity was associated with multiple visual and auditory markers including *speech prevalence* (*β* = −0.68; *P* =.017, *r*^2^ =.40, *overall expressivity* (*β* = −0.46; *P* = 0.10, *r*^2^ =.27), and head movement measured as *head pitch variability* (*β* = −1.24; *P* =.006, *r*^2^ =.48) and *head yaw variability* (*β* = −0.54; *p* =.055, *r*^2^ =.32).

**Conclusions:** Digital measurements of facial affect, movement, and speech prevalence demonstrated strong effect sizes and significant linear associations with severity of suicidal ideation.

## Introduction

Timely, efficient, sensitive, and non-invasive measurement approaches are required to improve suicide risk assessment [1]. One promising avenue is the use of remote digital monitoring methodologies that utilize smart devices such as phones and wearables in conjunction with deep learning algorithms to infer behavioral and physiological states associated with suicide risk [2,3].

While suicide risk is often comorbid with other mental disorders––in particular Major Depressive Disorder (MDD) and schizophrenia––suicidal behavior is increasingly recognized as unique in presentation and risk profile [4–7]. Based on prior knowledge, visual and auditory data sources represent a compelling direction in objective measurement of behavior associated with suicide risk. Emil Kraepelin first obserserved that suicide risk was associated with melancholic states characterized by slowed speech, where patients appear to “become mute in the middle of a sentence,” further observing how “the facial expression and the general attitude are sleepy and languid, the speech is low…” [8]. More contemporarily, reduced facial expressivity and movement measured using standardized coding schemes based on videos of patient interviews differentiated depressed patients with and without suicide risk [9] and altered vocal characteristics have been observed in acutely suicidal patients [5].

A number of visual and auditory characteristics can be directly quantified including gross motor activity [10], head movement variability [11–13], facial activity [14], and properties of speech [15]. The automated measurement of these clinical features introduces the possibility of objective digital assessment of visual and auditory markers of suicide risk. Given that audio and video data sources can be captured remotely, this further introduces the possibility of greatly scaling the reach and frequency of assessment. Increased scale and objectivity can facilitate increased accuracy and accessibility of clinical risk and treatment response assessment.

Visual and auditory biomarkers were selected based on a mechanistic theory that reduced serotonin––a key risk factor for suicidal behavior and a primary biological target for treatment of MDD––will affect behavioral characteristics including an individual’s speech, head movement, and facial activity. Serotonergic tone is known to mechanistically impact motor functioning directly and via interactions with dopamine and norepinephrine signalling [16–18]. Depleted serotonin has been observed in post-mortum brains of individuals with MDD [19] and those who have committed suicide [20]. Furthermore, direct pharmacological manipulation of serotonin using Selective Serotonin Reuptake Inhibitors (SSRIs) increases suicide risk [21,22].

While novel measurements are promising, validation is required before such metrics can be interpreted clinically. Key steps to validation include comparison to traditional clinical measures, both cross-sectionally and as they change alongside treatment and disease course [23]. Such measures should strive to be easy to collect, have increased sensitivity to facilitate frequent and accurate assessment, and should be validated in relationship to narrower biological phenotypes and treatment targets than traditional endpoints offer. This will lead to improved dynamic treatment research and clinical decision making [24] based on modulation of underlying neurobiological deficits [25].

In the current study, we examine measurements extracted from video interviews using open source deep learning algorithms to quantify facial, vocal, and movement behavior in relation to suicide risk severity in patients interviewed soon after admission to a psychiatric hospital following a suicide attempt.

## Methods

### Study Participants

Participants were recruited from the Psychiatric University Hospital, Zurich, Switzerland, as part of the “SIMON – Suicide Ideation MONitoring” study. The SIMON study is funded by the Digital Lives initiative of the Swiss National Science Foundation (SNSF) and was approved by the local ethical review board. It aims to develop a digital protocol to monitor and predict suicidal ideation and hospital readmission in high-risk psychiatric patients. Digital markers of psychopathology are set to be derived from smartphone-based experience sampling, mobile passive sensing, and video recordings of patients’ free speech.

Patients were included in the study if they met the following criteria: (i) admission to the hospital after a suicide attempt or in the context of suicidal ideation, and or suicidal ideation were identified in the first diagnostic intake interview, (ii) sufficient knowledge of the German language, (iii) being a smartphone user, (iv) discharge in accord with a clinician, with established outpatient care contact to the physician or psychologist. Patients were excluded if they met the following criteria: (i) planning to leave the greater Zurich area within the study period, (ii) sharing a smartphone with another person, (iv) being active military personnel. Researchers kept track of all patients admitted to the hospital and contacted the treating psychologist or physician in case of eligibility. Patients who met the inclusion criteria and for whom an approval from the psychologist or physician was granted were informed about the study. If patients agreed to participate in the study, they were invited for a baseline assessment appointment which entailed: (i) detailed information about the study and informed consent, (ii) assessment of current mental disorders with the Mini International Neuropsychiatric Interview (MINI; version 6), (iii) upon agreement – a short videotaped semi-structured qualitative interview, (iv) electronic/pen and paper questionnaires that evaluate relevant psychological variables, (v) smartphone apps installation. Participants were reimbursed with up to CHF 120.

Participants were recruited for the study on a rolling basis. At the time of the analysis, 30 patients agreed to participate in the videotaped interview, of whom 20 provided the necessary baseline questionnaires. Ultimately, this analysis was conducted on a sample size of 20 patients.

### Clinical assessment of suicidality

Assessment of suicide risk, was completed at baseline. The Beck Scale for Suicide Ideation questionnaire (BSS; German validated version) [26] was administered to evaluate patients’ current intensity of attitudes, plans, and behaviors to commit suicide. Patients’ history of non-suicidal self-injury (NSSI) and suicide attempts was assessed using two self-report items from the Self-Injurious Thoughts and Behaviors Interview (SITBI): (1) “In your life, have you purposefully hurt yourself without wanting to die?” and (2) “How many times in your lifetime have you made an attempt to kill yourself during which you had at least some intent to die?” [27,28].

### Collection of video and audio data

At baseline, patients were given the choice of additionally participating in a video-recorded interview. Upon agreement, patients signed an informed consent specific to this part of the study. A short videotaped semi-structured qualitative interview was performed. A laptop was placed on the table in front of the patient, and one-minute video & audio samples were recorded. During the qualitative video interview, participants answered introductory questions assessing their current state, and questions about experiences with different valences (i.e., neutral, positive, negative) and temporal dimensions (i.e., past, present, future). Altogether, six video and audio samples, each at one minute, were recorded per participant: introduction (neutral present), neutral, positive past, positive future, negative past, negative future. Supplementary Table 1 displays exemplary questions asked for each category. The conversation for each category starts with the central question (first in order) asked by the researcher, followed by additional questions to keep the participant involved and talking during the one-minute recordings.

### Machine learning computer vision and voice analysis

All analysis was conducted in a Python environment using open source tools. All code used to calculate visual and auditory biomarkers, the resulting data output, and the executable python script for the analysis of the data output as is presented in this manuscript is available on Github [35]. Analyses were conducted on the entire video interview.

### Facial Activity Analysis

First, all videos were segmented into frames resulting in a minimum of 33 image frames per second. Next, each image was segmented into three matrices consisting of red, blue, green spectrum pixels for use in computer vision (CV) modeling using OpenCV, a open source computer vision software package [29]. Subsequently, each frame was analysed using OpenFace [30], an open source software package that has demonstrated validity next to expert human ratings of Facial Action Coding Scheme (FACS) [31], a standardized methodology to measure facial movements that reflect activity in the underlying human facial musculature utilized in the production of basic emotions. Specifically, for each frame, OpenFace outputs (i) a binary value indicating the presence of an action unit and (ii) a continuous value indicating the intensity with which the action unit is being expressed. Action unit intensities were used to derive measures of individual emotional expressivity i.e. *happiness expressivity, sadness expressivity, fear expressivity, anger expressivity, surprise expressivity*, and *disgust expressivity*. Action unit intensities were also used to calculate *overall expressivity* regardless of emotion.

### Movement Analysis

The angle of the head’s pitch (up and down movement) and yaw (side to side movement) was acquired for each video frame using OpenFace. The standard deviation of the framewise pitch and yaw measurements were used as indicators of head movement i.e. *head pitch variability* and *head yaw variability*.

### Voice analysis

Recordings were segmented into speech and non-speech parts using Parselmouth, an open source software package utilized for vocal analysis [32]. The percentage of the audio file where speech was recorded as opposed to no speech i.e. *speech prevalence* was calculated to represent how much the participant was talking given the length of the recording.

### Data Analysis

First, BSS scores were regressed on all movement, facial, and audio markers controlling for age and gender using separate multiple linear regressions. In addition to significance testing, unique variance accounted for in BSS scores by the digital measures was assessed using Cohen’s *d* [33]. Following analysis of *overall expressivity*, the BSS was regressed on expressivity of each emotion, correcting for multiple comparisons using false discovery rate adjustment to reduce *P* value inflation due to chance.

## Results

### Suicide Severity

Suicidal ideation severity on the BSS across subjects was, on average, above the clinical cut-off of 9 indicating severe suicide risk (*μ* = 10.9; *σ* = 10.2; range = 0-31) and an average count of 3 past suicide attempts (*σ* = 6.9; range 0-30).

### Facial Activity

Controlling for age and gender, *overall expressivity* demonstrated a significant negative linear association with BSS scores, indicating decreased facial activity is associated with greater suicide risk (*β* = −0.46; *P* = 0.10, *r*^2^ =.27). This indicates that facial activity decreases as suicide severity increases, with the strongest evidence in the context of facial expressions without emotional expressions.

To better understand if particular emotions contributed to the observation of *composite expressivity*, we regressed BSS scores with individual emotional expressivity, gender, and age. P-values were adjusted using false discovery rate adjustment. Post hoc-analyses demonstrated significance using Benjamini-Hochberg Adjusted *p*-value, for *sadness expressivity* (*β* = - 0.68; *P* =.013, *r*^2^ =.43), *surprise expressivity* (*β* = - 0.74.; *P =*.002, *r*^2^ =.53), and *disgust expressivity* (*β* = - 0.64.; *P =*.*036, r*^2^ =.35), but not for *fear, anger, happiness expressivity* (Table 1).

**Table 1:** Results from multiple linear regression between BSS scores and digital measurements of facial expressivity, vocal behavior, and head movement, controlling for age and gender.

| Predictor | Coefficient | Std. Error | t | p-value | F | R<sup>2</sup> | Adj. R<sup>2</sup> | p-value |
| --- | --- | --- | --- | --- | --- | --- | --- | --- |
| Constant | 0.2020 | 0.380 | 0.531 | 0.603 | 0.9013 | 0.153 | -0.017 | 0.464 |
| **Anger expressivity** | -0.2456 | 0.322 | -0.762 | 0.458 |  |  |  |  |
| Age | 0.0068 | 0.007 | 0.960 | 0.352 |  |  |  |  |
| Gender | 0.1167 | 0.172 | 0.680 | 0.507 |  |  |  |  |
| Constant | 0.1213 | 0.240 | 0.506 | 0.620 | 2.693 | 0.350 | 0.220 | 0.0834 |
| **Disgust expressivity** | -0.6401 | 0.278 | -2.305 | 0.036 |  |  |  |  |
| Age | 0.0120 | 0.006 | 1.980 | 0.066 |  |  |  |  |
| Gender | 0.1422 | 0.146 | 0.972 | 0.346 |  |  |  |  |
| Constant | 0.2628 | 0.380 | 0.692 | 0.499 | 1.043 | 0.173 | 0.007 | 0.402 |
| **Fear expressivity** | -0.3210 | 0.328 | -0.978 | 0.344 |  |  |  |  |
| Age | 0.0060 | 0.007 | 0.842 | 0.413 |  |  |  |  |
| Gender | 0.1056 | 0.170 | 0.620 | 0.544 |  |  |  |  |
| Constant | 0.0051 | 0.292 | 0.018 | 0.986 | 0.6834 | 0.120 | -0.056 | 0.575 |
| **Happiness expressivity** | -0.0228 | 0.255 | -0.089 | 0.930 |  |  |  |  |
| Age | 0.0083 | 0.007 | 1.194 | 0.251 |  |  |  |  |
| Gender | 0.1517 | 0.178 | 0.852 | 0.408 |  |  |  |  |
| Constant | 0.4394 | 0.270 | 1.629 | 0.124 | 3.714 | 0.426 | 0.311 | 0.0353 |
| **Sadness expressivity** | -0.6785 | 0.240 | -2.830 | 0.013 |  |  |  |  |
| Age | 0.0082 | 0.006 | 1.480 | 0.160 |  |  |  |  |
| Gender | 0.0345 | 0.152 | -0.228 | 0.823 |  |  |  |  |
| Constant | -0.0441 | 0.198 | -0.223 | 0.827 | 5.713 | 0.533 | 0.440 | 0.00819 |
| **Surprise expressivity** | -0.7437 | 0.204 | -3.645 | 0.002 |  |  |  |  |
| Age | 0.0143 | 0.005 | 2.734 | 0.015 |  |  |  |  |
| Gender | 0.2421 | 0.127 | 1.912 | 0.075 |  |  |  |  |
| Constant | 0.2881 | 0.300 | 0.961 | 0.352 | 1.820 | 0.267 | 0.120 | 0.187 |
| **Overall expressivity** | -0.4585 | 0.264 | -1.734 | 0.103 |  |  |  |  |
| Age | 0.0054 | 0.006 | 0.833 | 0.418 |  |  |  |  |
| Gender | 0.1947 | 0.158 | 1.234 | 0.236 |  |  |  |  |
| Constant | 0.3291 | 0.256 | 1.285 | 0.218 | 3.389 | 0.404 | 0.285 | 0.0459 |
| **Speech prevalence** | -0.6808 | 0.255 | -2.674 | 0.017 |  |  |  |  |
| Age | 0.0057 | 0.006 | 0.991 | 0.337 |  |  |  |  |
| Gender | 0.3429 | 0.158 | 2.170 | 0.047 |  |  |  |  |
| Constant | 0.4192 | 0.248 | 1.689 | 0.112 | 4.533 | 0.476 | 0.371 | 0.018 |
| **Head pitch variability** | -1.2465 | 0.391 | -3.189 | 0.006 |  |  |  |  |
| Age | 0.0124 | 0.005 | 2.294 | 0.037 |  |  |  |  |
| Gender | -0.0342 | 0.143 | -0.239 | 0.814 |  |  |  |  |
| Constant | 0.1006 | 0.245 | 0.411 | 0.687 | 0.117 | 0.317 | 0.181 | 0.117 |
| **Head yaw variability** | -0.5408 | 0.260 | -2.081 | 0.055 |  |  |  |  |
| Age | 0.0125 | 0.006 | 1.979 | 0.066 |  |  |  |  |
| Gender | 0.1723 | 0.150 | 1.146 | 0.270 |  |  |  |  |

### Voice Analysis

Controlling for age and gender, *speech prevalence* demonstrated a significant negative linear association with BSS scores, indicating that suicide severity scores increased as speech decreased (*β* = −0.68; *P* =.017, *r*^2^ =.40; Table 1).

### Movement Analysis

Controlling for age and gender, both *head pitch variability* and *head yaw variability* demonstrated significant negative linear relationships with suicide severity (*β*_*pitch*_ = −1.24; *P* =.006, *r*^2^ =.48; *β*_*yaw*_ = −0.54; *p* =.055, *r*^2^ =.32), indicating that lower levels of head movement were associated with greater suicide severity scores (Table 1).

## Discussion

We examined visual and auditory measures of facial activity, head movement, and speech production, all calculated using deep learning algorithms applied to open-ended clinical interviews with psychiatric patients following a suicide attempt. The goal of this work was to determine if key indicators of the suicide severity could be measured in an objective and automated manner using video data captured during clinical interviews that provided structured questions, but were otherwise kept deliberately open to mimick psychiatric interviewing in routine care. Achieving this goal provides a proof-of-concept that suicide risk can be assessed through analysis of unstructured video interview data in conjunction with deep-learning algorithms designed to measure clinical/behavioral characteristics.

Direct, objective measurement of visual and auditory markers of suicide severity can be built in to any context where video and audio of patients is being transmitted. The current study did not use any specialized recording equipment; rather a commercially available tablet was used to record video. Such models, for example, can be built into the back-end of any secure tele-health communication. As both public and private telehealth systems rapidly become more ubiquitous, there is a need to incorporate objective measurement to identify and communicate clinical risk. The benefit of using unstructured clinical conversations is that such markers can be measured during any conversation, thus providing non-invasive, passive measurement of suicide severity. Ultimately, methods that can provide remote, objective, and passive measurement of clinical functioning can greatly increase the reach of treatment development, dissemination, and personalization [24,34].

Finally, the markers that were studied were selected based on a mechanistic theory that reduced serotonin, a key risk factor for suicidal behavior, will affect motor and movement characteristics including patient’s speech, head movement, and facial activity. These primary hypotheses were confirmed. This work provides a proof-of-concept that proxy markers of underlying neurobiological functioning can be used to measure clinical risk. Demonstrating this indicates that there is potential to remotely measure more specific neurobiological phenotypes to determine risk or examine the response to treatment. For example, due to suicide risk associated with SSRIs, there is a need for close clinical monitoring in during the initiation of treatment. Similarly, such models may be useful to determine who would benefit from treatments that affect serotonergic tone. Ultimately, by measuring a more specific, biologically based phenotype, there is greatly improved opportunity to improve sensitivity of measurement and specificity of treatment [25].

The work presents with key limitations. Firstly, the current work represents a proof-of-concept. The use of such markers for risk assessment or measurement of clinical severity should only be done in an exploratory manner until such markers can be validated in larger and more diverse clinical populations and settings. It should be directly determined the amount of video data needed and clinically meaningful cutoffs to determine clinical functioning. It is likely that, like traditional suicide risk assessment, multiple characteristics together, are needed to determine risk or severity. Multiple digital measures can be combined to produce a more robust metric, but like traditional scale development, this requires larger samples from diverse populations. Importantly, the current work uses open-source software and additionally, we publish our software methods. As such, there is no barrier for researchers to access to implement the identical methods to test and extend the current approach. An open-source approach lends itself to rapid replication, extension, and implementation.

Ultimately, this proof-of-concept study demonstrates that theoretically grounded visual and auditory digital measurements demonstrate validity as markers of suicide severity. This effort provided a digital approach akin to traditional clinical assessment whereby a skilled clinician listens to a patient and applies internal working models developed through years of experience to assess clinical functioning.

## Data Availability

Due to privacy concerns, participants of this study did not agree for their data to be shared publicly.

## Supplementary Materials

**Supplementary Table 1:**
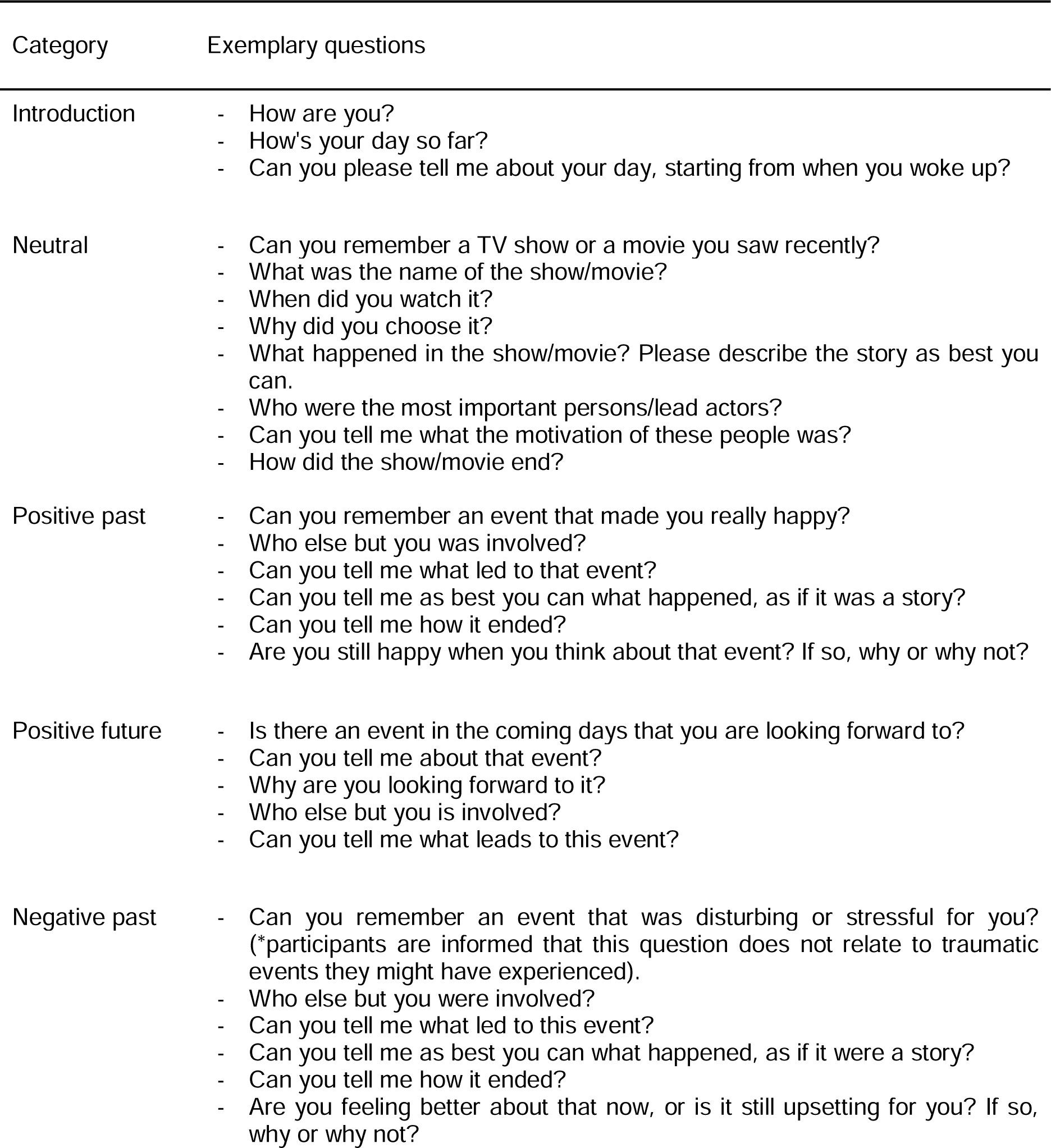
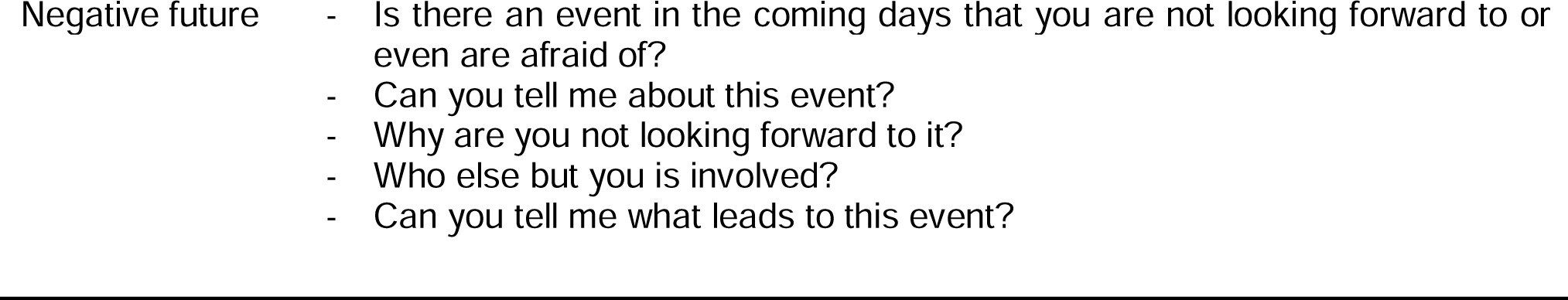
Exemplary questions for the six categories of the videotaped, semi-structured qualitative interview.

| Category | Exemplary questions |
| --- | --- |
| Introduction | - How are you?<br>- How's your day so far?<br>- Can you please tell me about your day, starting from when you woke up? |
| Neutral | - Can you remember a TV show or a movie you saw recently?<br>- What was the name of the show/movie?<br>- When did you watch it?<br>- Why did you choose it?<br>- What happened in the show/movie? Please describe the story as best you can.<br>- Who were the most important persons/lead actors?<br>- Can you tell me what the motivation of these people was?<br>- How did the show/movie end? |
| Positive past | - Can you remember an event that made you really happy?<br>- Who else but you was involved?<br>- Can you tell me what led to that event?<br>- Can you tell me as best you can what happened, as if it was a story?<br>- Can you tell me how it ended?<br>- Are you still happy when you think about that event? If so, why or why not? |
| Positive future | - Is there an event in the coming days that you are looking forward to?<br>- Can you tell me about that event?<br>- Why are you looking forward to it?<br>- Who else but you is involved?<br>- Can you tell me what leads to this event? |
| Negative past | - Can you remember an event that was disturbing or stressful for you? (*participants are informed that this question does not relate to traumatic events they might have experienced).<br>- Who else but you were involved?<br>- Can you tell me what led to this event?<br>- Can you tell me as best you can what happened, as if it were a story?<br>- Can you tell me how it ended?<br>- Are you feeling better about that now, or is it still upsetting for you? If so, why or why not? |

## References

1. The Agenda Development Process of the United States’ National Action Alliance for Suicide Prevention Research Prioritization Task Force. Crisis Hogrefe Publishing; 2013 Apr 29;34(3):147–155. [doi: 10.1027/0227-5910/a000208]

2. Torous J, Walker R. Leveraging Digital Health and Machine Learning Toward Reducing Suicide-From Panacea to Practical Tool. JAMA Psychiatry 2019 Jul 10; PMID:31290952

3. Vahabzadeh A, Sahin N, Kalali A. Digital Suicide Prevention: Can Technology Become a Game-changer? Innov Clin Neurosci 2016 Jun 1;13(5–6):16–20. PMID:27800282

4. American Psychiatric Association. Diagnostic and Statistical Manual of Mental Disorders [Internet]. Fifth Edition. American Psychiatric Association; 2013 [cited 2020 Aug 31]. [doi: 10.1176/appi.books.9780890425596] ISBN:978-0-89042-555-8

5. Cummins N, Scherer S, Krajewski J, Schnieder S, Epps J, Quatieri TF. A review of depression and suicide risk assessment using speech analysis. Speech Commun 2015 Jul 1;71:10–49. [doi: 10.1016/j.specom.2015.03.004]

6. Pompili M, Amador XF, Girardi P, Harkavy-Friedman J, Harrow M, Kaplan K, Krausz M, Lester D, Meltzer HY, Modestin J, Montross LP, Bo Mortensen P, Munk-Jørgensen P, Nielsen J, Nordentoft M, Saarinen PI, Zisook S, Wilson ST, Tatarelli R. Suicide risk in schizophrenia: learning from the past to change the future. Ann Gen Psychiatry 2007 Mar 16;6(1):10. [doi: 10.1186/1744-859X-6-10]

7. Sisti D, Mann JJ, Oquendo MA. Toward a Distinct Mental Disorder-Suicidal Behavior. JAMA Psychiatry 2020 Mar 18; PMID:32186693

8. Kraepelin E. Clinical psychiatry. Macmillan; 1907.

9. Heller M, Haynal V. Depression and Suicide Faces. What Face Reveals Basic Appl Stud Spontaneous Expr Using Facial Action Coding Syst FACS 2005.

10. Buyukdura JS, McClintock SM, Croarkin PE. Psychomotor retardation in depression: biological underpinnings, measurement, and treatment. Prog Neuropsychopharmacol Biol Psychiatry 2011 Mar 30;35(2):395–409. PMID:21044654

11. Alghowinem S, Goecke R, Wagner M, Parkerx G, Breakspear M. Head Pose and Movement Analysis as an Indicator of Depression. Proc 2013 Hum Assoc Conf Affect Comput Intell Interact [Internet] USA: IEEE Computer Society; 2013 [cited 2020 Aug 31]. p. 283–288. [doi: 10.1109/ACII.2013.53]

12. Dibeklioğlu H, Hammal Z, Cohn JF. Dynamic Multimodal Measurement of Depression Severity Using Deep Autoencoding. IEEE J Biomed Health Inform 2018 Mar;22(2):525–536. [doi: 10.1109/JBHI.2017.2676878]

13. Kim Y, Cheon S-M, Youm C, Son M, Kim JW. Depression and posture in patients with Parkinson’s disease. Gait Posture 2018;61:81–85. PMID:29306811

14. Girard JM, Cohn JF, Mahoor MH, Mavadati S, Rosenwald DP. Social Risk and Depression: Evidence from Manual and Automatic Facial Expression Analysis. Proc Int Conf Autom Face Gesture Recognit IEEE Int Conf Autom Face Gesture Recognit 2013;1–8. PMID:24598859

15. Garcia-Toro M, Talavera JA, Saiz-Ruiz J, Gonzalez A. Prosody Impairment in Depression Measured through Acoustic Analysis. J Nerv Ment Dis 2000 Dec;188(12):824–829.

16. El Mansari M, Guiard BP, Chernoloz O, Ghanbari R, Katz N, Blier P. Relevance of Norepinephrine–Dopamine Interactions in the Treatment of Major Depressive Disorder. CNS Neurosci Ther 2010 Apr 8;16(3):e1–e17. PMID:20406250

17. Herrera-Guzmán I, Gudayol-Ferré E, Herrera-Guzmán D, Guàrdia-Olmos J, Hinojosa-Calvo E, Herrera-Abarca JE. Effects of selective serotonin reuptake and dual serotonergic-noradrenergic reuptake treatments on memory and mental processing speed in patients with major depressive disorder. J Psychiatr Res 2009 Jun;43(9):855–863. PMID:19128810

18. Morrissette DA, Stahl SM. Modulating the serotonin system in the treatment of major depressive disorder. CNS Spectr 2014 Dec;19 Suppl 1:57–67; quiz 54–57, 68. PMID:25544378

19. Stockmeier CA. Involvement of serotonin in depression: evidence from postmortem and imaging studies of serotonin receptors and the serotonin transporter. J Psychiatr Res 2003 Oct;37(5):357–373. PMID:12849929

20. Stockmeier CA, Shapiro LA, Dilley GE, Kolli TN, Friedman L, Rajkowska G. Increase in Serotonin-1A Autoreceptors in the Midbrain of Suicide Victims with Major Depression— Postmortem Evidence for Decreased Serotonin Activity. J Neurosci Society for Neuroscience; 1998 Sep 15;18(18):7394–7401. PMID:9736659

21. Lancet T. SSRIs: suicide risk and withdrawal. The Lancet Elsevier; 2003 Jun 14;361(9374):1999. PMID:12814704

22. Wessely S, Kerwin R. Suicide Risk and the SSRIs. JAMA American Medical Association; 2004 Jul 21;292(3):379–381. [doi: 10.1001/jama.292.3.379]

23. Coravos A, Khozin S, Mandl KD. Developing and adopting safe and effective digital biomarkers to improve patient outcomes. Npj Digit Med Nature Publishing Group; 2019 Mar 11;2(1):1–5. [doi: 10.1038/s41746-019-0090-4]

24. Torous J, Onnela J-P, Keshavan M. New dimensions and new tools to realize the potential of RDoC: digital phenotyping via smartphones and connected devices. Transl Psychiatry 2017 07;7(3):e1053. PMID:28267146

25. Lenze EJ, Rodebaugh TL, Nicol GE. A Framework for Advancing Precision Medicine in Clinical Trials for Mental Disorders. JAMA Psychiatry 2020 Mar 25; PMID:32211837

26. Kliem S, Lohmann A, Mößle T, Brähler E. German Beck Scale for Suicide Ideation (BSS): psychometric properties from a representative population survey. BMC Psychiatry 2017 Dec 4;17(1):389. [doi: 10.1186/s12888-017-1559-9]

27. Nock MK, Holmberg EB, Photos VI, Michel BD. Self-Injurious Thoughts and Behaviors Interview: development, reliability, and validity in an adolescent sample. Psychol Assess 2007 Sep;19(3):309–317. PMID:17845122

28. Chu C, Hom MA, Stanley IH, Gai A, Nock MK, Gutierrez PM, Joiner TE. Non-Suicidal Self-Injury and Suicidal Thoughts and Behaviors: A Study of the Explanatory Roles of the Interpersonal Theory Variables among Military Service Members and Veterans. J Consult Clin Psychol 2018 Jan;86(1):56–68. PMID:29172592

29. Brahmbhatt S. Introduction to Computer Vision and OpenCV. Pract OpenCV [Internet] Berkeley, CA: Apress; 2013 [cited 2020 Sep 1]. p. 3–5. [doi: 10.1007/978-1-4302-6080-6_1]

30. Baltrusaitis T, Robinson P, Morency L-P. OpenFace: An open source facial behavior analysis toolkit. 2016 IEEE Winter Conf Appl Comput Vis WACV [Internet] Lake Placid, NY, USA: IEEE; 2016 [cited 2020 May 29]. p. 1–10. [doi: 10.1109/WACV.2016.7477553]

31. Ekman P, Rosenberg EL. What the Face RevealsBasic and Applied Studies of Spontaneous Expression Using the Facial Action Coding System (FACS) [Internet]. Oxford University Press; 2005 [cited 2020 Oct 9]. [doi: 10.1093/acprof:oso/9780195179644.001.0001] ISBN: 978-0-19-517964-4

32. Jadoul Y, Thompson B, de Boer B. Introducing Parselmouth: A Python interface to Praat. J Phon 2018 Nov;71:1–15. [doi: 10.1016/j.wocn.2018.07.001]

33. Cohen ML. An Adaptive R-Estimate: [Internet]. Fort Belvoir, VA: Defense Technical Information Center; 1980 Dec. [doi: 10.21236/ADA096768]

34. Insel TR. Digital phenotyping: a global tool for psychiatry. World Psychiatry 2018 Oct;17(3):276–277. PMID:30192103

35. Abbas A. anzarabbas/ms_dbms_mdd_suicidality [Internet]. 2020 [cited 2020 Oct 21]. Available from: https://github.com/anzarabbas/ms_dbms_mdd_suicidality

